# Evaluation of pooling of samples for testing SARS-COV- 2 for mass screening of COVID-19

**DOI:** 10.1101/2021.03.15.21253567

**Authors:** Sally Mahmoud, Esra Ibrahim, Bhagyashree Thakre, Juliet Teddy, Preety Raheja, Subhashini Ganesan, Walid Zaher

**Affiliations:** Biogenix G42 lab, Abu Dhabi, UAE; G42 Health care, Abu Dhabi, UAE

**Keywords:** COVID-19, SARS-CoV-2, diagnosis, mass screening, sample pooling

## Abstract

**Background:** The current pandemic of SARS- COV- 2 virus, widely known as COVID-19 has affected millions of people around the world. The World Health Organization (WHO) has recommended vigorous testing to differentiate SARS-CoV-2 from other respiratory infections to aid in guiding appropriate care and management. Situations like this have demanded robust testing strategies and pooled testing of samples for SARS- COV- 2 virus has provided the solution to mass screening of people. The pooled testing strategy can be very effective in testing with limited resources, yet it comes with its own limitations. These limitations need critical consideration when it comes to testing of highly infectious disease like COVID −19.

**Methods:** The study evaluated the pooled testing of nasopharyngeal swabs for SARS- COV- 2 by comparing sensitivity of individual sample testing with 4 and 8 pool sample testing. Median cycle threshold (Ct) values were compared. The precision of pooled testing was assessed by doing an inter and intra assay of pooled samples. Coefficient of variance was calculated for inter and intra assay variability.

**Results:** The sensitivity becomes considerably low when the samples are pooled, there is a higher percentage of false negatives with higher pool size and when the patient viral load is low or weak positive samples. High variability was seen in the intra and inter assay, especially in weak positive samples and larger pool size.

**Conclusion:** As COVID - 19 numbers are still high and testing capacity needs to be high, we have to meticulously evaluate the testing strategy for each country depending on its testing capacity, infrastructure, economic strength, and need to make a serious call on cost effective strategy of resource saving and risk/ cost of missing positive patients.

## Background

The world in 2020 has been witnessing the most crippling pandemic on earth, ever since its outbreak in December 2019 at Wuhan, China. It is widely known as COVID-19, caused by SARS-COV-2 virus, it’s a new virus and not much has been known about the virus. From what is known the virus seems to exhibit a variable incubation period which can be up to 14 days and asymptomatic carriers can also transmit the virus, it spreads rapidly and affected large number of people than SARS and MERS, despite having lower case fatality rate. ^1^ All these characteristics of the novel virus has made the containment and control of the virus challenging.

The World Health Organization (WHO) has recommended robust diagnostic testing to differentiate SARS-CoV-2 from other respiratory infections to aid in guiding appropriate care and management. Since the pandemic has affected millions of people, mass testing requires a lot of resources. Further the WHO has suggested around 10 – 30 tests per Positive case as a benchmark for adequate testing, it also has recommended a positive rate lower than 10% and better if lower than 3%. ^2^

In situations like this sample pooling can be a vital strategy to do testing in large numbers, where sample extracts from a random number of samples from a heterogeneous population group are combined into a single tube for pooled PCR analysis and this strategy have shown to be cost–effective during mass testing compared with individual testing. ^3^

When the disease prevalence is less, it can be advantageous to pool individual samples into a single pool, this increases the test capacity and reduces the number of PCR tests. ^4,5^ For COVID-19, it has been estimated that pooling strategy reduces cost by 69% and use of tenfold fewer tests. ^6,7^ Recent research have also established the optimal pool size that maintains the testing accuracy for SARS-CoV-2 PCR and has found that accuracy is retained in a pool size of up to 32 samples. It has shown that costs can be reduced substantially without sacrificing accuracy. ^8-10^

However, when samples are diluted, there could be less viral genetic material available to detect and this increases in greater likelihood of false negative results. But studies establish that sample pooling will greatly increase the number of individuals that can be tested using less resource with a small reduction in sensitivity, which may be acceptable depending on the pooling efficiency ^11^ Therefore, the FDA generally recommends that pooling test shows ≥85% percent positive agreement when compared with the same test performed on individual samples.

In UAE, as of November 2020, there are around 1500 COVID −19 cases per 100K population and UAE is in the top compared to other nations in the number of tests done per 100K population. ^12^ Around 149,000 tests are done per 100K population and the positive rate is 1%. [Figure 1]. With this background, the study aims to evaluate the pooling strategy for mass screening of COVID-19.

**Fig 1:**
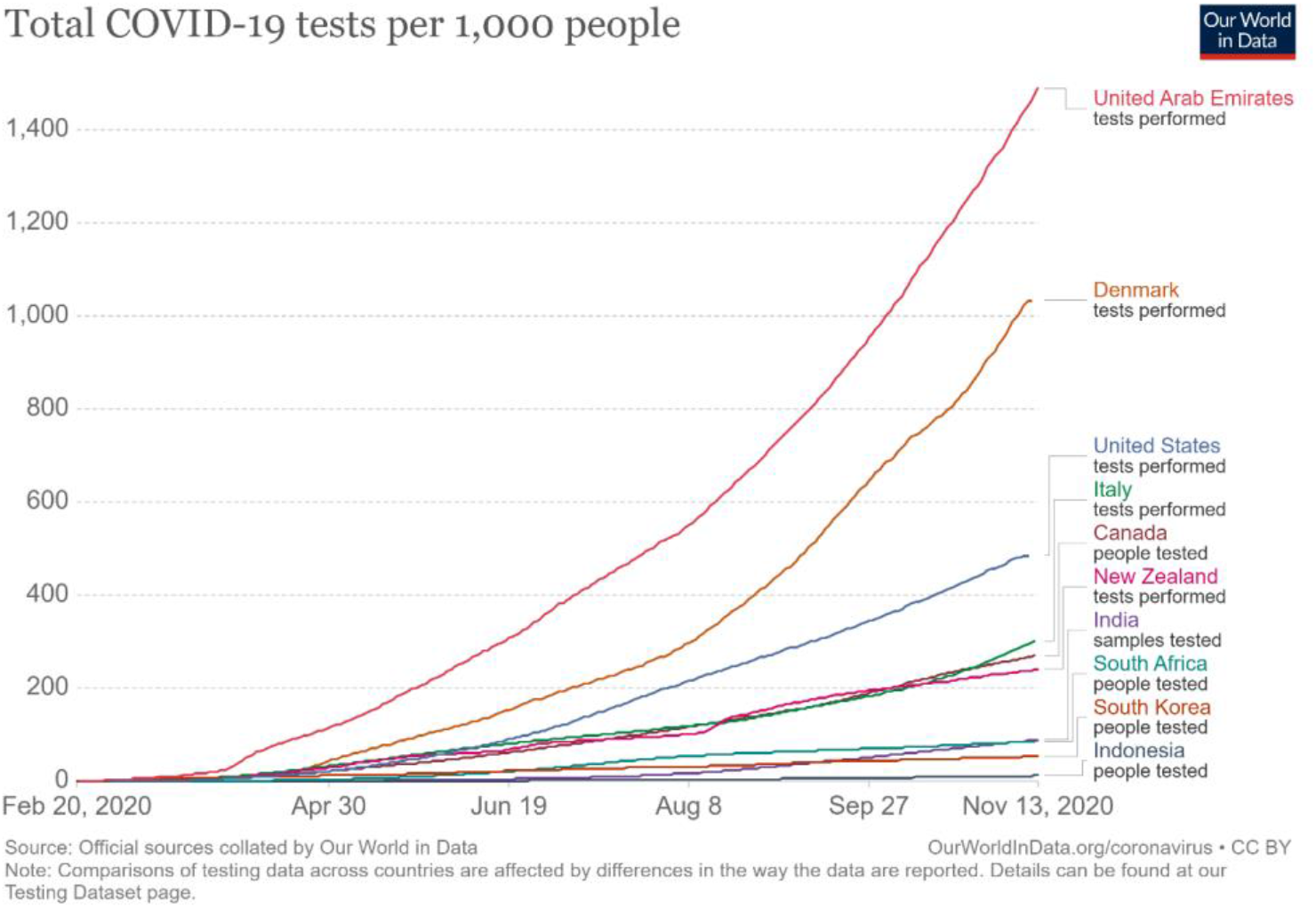
Shows the number of COVID – 19 tests done per 1000 people 13.

### Aim

The aim of the study is to evaluate the pooling method for mass screening of COVID-19.

### Objectives

- To evaluate the sensitivity of 4 and 8 sample pooling for testing COVID −19.
- To study the repeatability and reproducibility of sample pooling by doing a inter and intra assay precision analysis

## Materials and method

The samples for this study were collected from individuals using nasopharyngeal swabs and were transported to the viral transport medium (VTM). All methods were carried out in accordance with relevant guidelines and regulations. Then RNA extraction was done by the automated machine MGISP −960. After the RNA extraction, 10 microliters of the sample extract is added to 20 microliter of the master mix (BGI RT-PCR fluorescence KIT). Every plate has a positive control and a negative or blank control.

In individual sample extraction, 160 microliters of the sample were used in each well, for the pooled sample 160 microlitre in each well was constituted by 40 microlitre of each sample in 4 fold pool sampling and 20 microlitre of each sample for 8 fold pool sampling.

Then the pooled samples are extracted following the RT-PCR procedure. VIC channel represents the B-actin housekeeping gene as internal control while FAM channel represents the ORF1ab gene for SARS-COV-2 virus detection. A positive test specimen is one for which VIC must have a cycle threshold (Ct) value ≤ 32 and FAM Ct ≤ 35, both fluorescence curves should be S shaped. A negative sample is one where there is no fluorescence curve at the FAM channel and a CT value of<32 and S shaped curve at the VIC channel.

If “S” shaped fluorescence curve is detected with a FAM Ct > 35, re-extraction is done and RT PCR test is repeated, if a negative result is detected, it is reported negative and if “S” curve is detected with a FAM Ct ≤ 38 in re extraction results, then a positive report is given.

### Sensitivity of pooled sample

For the sensitivity analysis, 40 known positive samples and 280 known negative samples were taken. In the 4-fold pool sample testing 40 known positive samples and 120 known negative samples were tested and in 8-fold pool sample testing, 40 known positive samples and 280 known negative samples were tested. Each well contained one known positive and 3 known negative samples in the 4pool method and each well contained one known positive and 7 known negative samples in the 8-pool sampling.

To analyse the sensitivity of sample of various viral load, out of the 40 samples used for pooling, ten high viral load samples [HP] with Ct≤20, ten medium viral load samples [MP] with Ct > 20 to <30 and twenty low viral load or weak positives samples [WP] with Ct > 30 were used.

The sensitivity of the 4-pool sampling and 8 - pool sampling was determined by running the samples individually and the same samples were run in 4 and 8 pooled sample method and the ability to detect the true positives were compared.

### Precision

To assess the precision of pooled sample method intra and inter assay precision study were conducted.

### Intra assay precision

Intra assay precision was calculated by running 30 known positive samples in 4 pool and 8 pool sampling twice. 30 known positive samples were classified into three groups of 10 samples each based on the CT values as high viral load samples [HP] medium viral load samples [MP] and low viral load or weak positives samples [WP] as mentioned above.

All these samples were run twice, and the variability was assessed calculating the coefficient of variation (CoV %).

### Inter assay precision

Inter assay precision was calculated by running 30 known positive samples, which was again classified into three groups based on the CT values as above. These samples were run on three consecutive days and the variability was assessed calculating the coefficient of variation (CoV %).

While doing the intra and inter assay experiment three known negative samples have been added to the same well along with one of the 30 known positive samples for 4 pool sample and seven known negative were added to the same pool along with one known positive for 8 pool sample.

## Results

Our study showed that the sensitivity of 4 pooled sample was 75% and the sensitivity of 8 pooled sample was 62.5% (Table 1 and table 2). The sensitivity decreases from 75% in 4 pool sampling to 62.5% in 8 pool sampling. In the low viral load, weak positive samples (Ct value ≥ 35), sensitivity of 4 pool sample was 50% (Table 3) and it further reduces to 25% in 8 pool sampling with a false negative percentage as high as 75%. (Table 4)

**Table 1:** Sensitivity of 4 sample pooling.

| Pooling method | Standard method (Individual sample run) |  |  |
| --- | --- | --- | --- |
|  | Positive | Negative | Total |
| Positive | 30 | 0 | 30 |
| Negative | 10 | 0 | 10 |
| Total | 40 |  | 40 |

Sensitivity– 75% (CI - 61.7% - 88.3%)
False negative percentage – 25%

**Table 2:** Sensitivity of 8 sample pooling.

| Pooling method | Standard method |  |  |
| --- | --- | --- | --- |
|  | Positive | Negative | Total |
| Positive | 25 | 0 | 27 |
| Negative | 15 | 0 | 13 |
| Total | 40 |  | 40 |

Sensitivity – 62.5% (CI – 47.5% - 77.5%)
False negative percentage – 37.5%

**Table 3:** **Sensitivity of 4 sample pooling of 20 weak positive samples with Ct value more than 35**

| Pooling method | Standard method |  |  |
| --- | --- | --- | --- |
|  | Positive | Negative | Total |
| Positive | 10 | 0 | 30 |
| Negative | 10 | 0 | 10 |
| Total | 20 |  | 40 |

Sensitivity– 50 % (CI – 28.09 % - 71.91%)
False negative percentage – 50%

**Table 4:** **Sensitivity of 8 sample pooling of 20 weak positive samples with Ct value more than 35**

| Pooling method | Standard method |  |  |
| --- | --- | --- | --- |
|  | Positive | Negative | Total |
| Positive | 5 | 0 | 5 |
| Negative | 15 | 0 | 15 |
| Total | 20 |  | 20 |

Sensitivity – 25 % ( CI – 6.03 % - 43.97%)
False negative percentage – 75%

The Ct difference was calculated between individual and pooled sample and the difference increased with the increase in pool size, the difference was more pronounced in the low viral load, weak positive group. (Table 5). The median Ct difference increased with increase in pool size and the maximum values were observed in weak viral load positive pooled samples. In the weak viral load positive sample pooling, the median became almost 0, as most samples turned negative with Ct value of 0 in the 8 pool of weak viral load positive sample. [Fig 2, 3, 4]

**Table 5:** Median Ct difference between individual and pooled samples.

| Pool details | Ct difference (Median) | Interquartile range (IQR) |
| --- | --- | --- |
| 4 pool (HP) | 2.29 | 0.49 |
| 4 pool (MP) | 1.79 | 0.79 |
| 4 pool (WP) | - 18.26 | 39.60 |
| 8 pool (HP) | 3.10 | 1.38 |
| 8 pool (MP) | 2.66 | 1.21 |
| 8 pool (WP) | -36.01 | 28.81 |

\* HP – High viral load positive sample, MP- Medium viral load positive sample, WP- low
viral load/ weak positive sample

**Figure 2:**
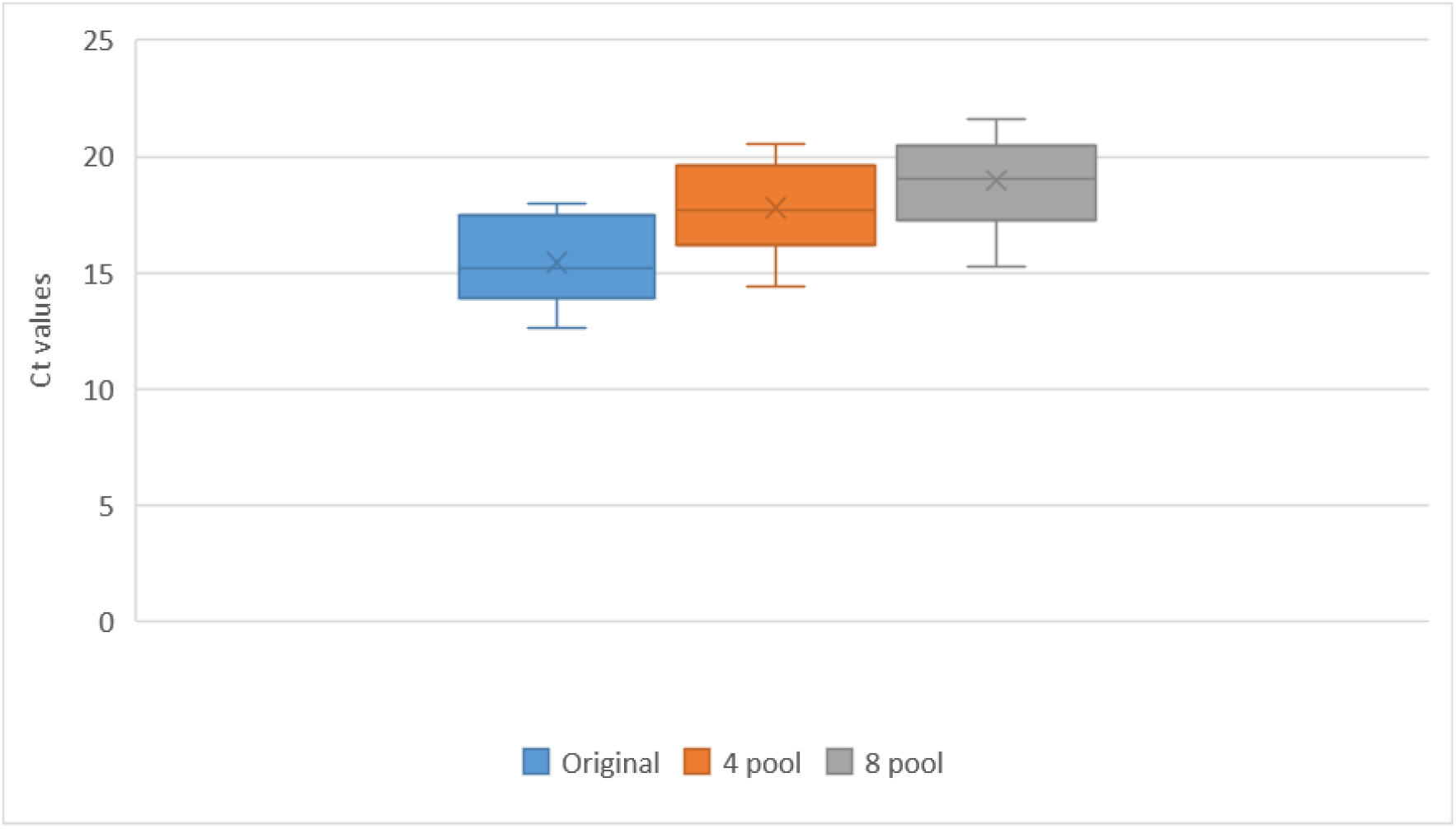
The Ct values of High viral load positive (HP) samples.

**Figure 3:**
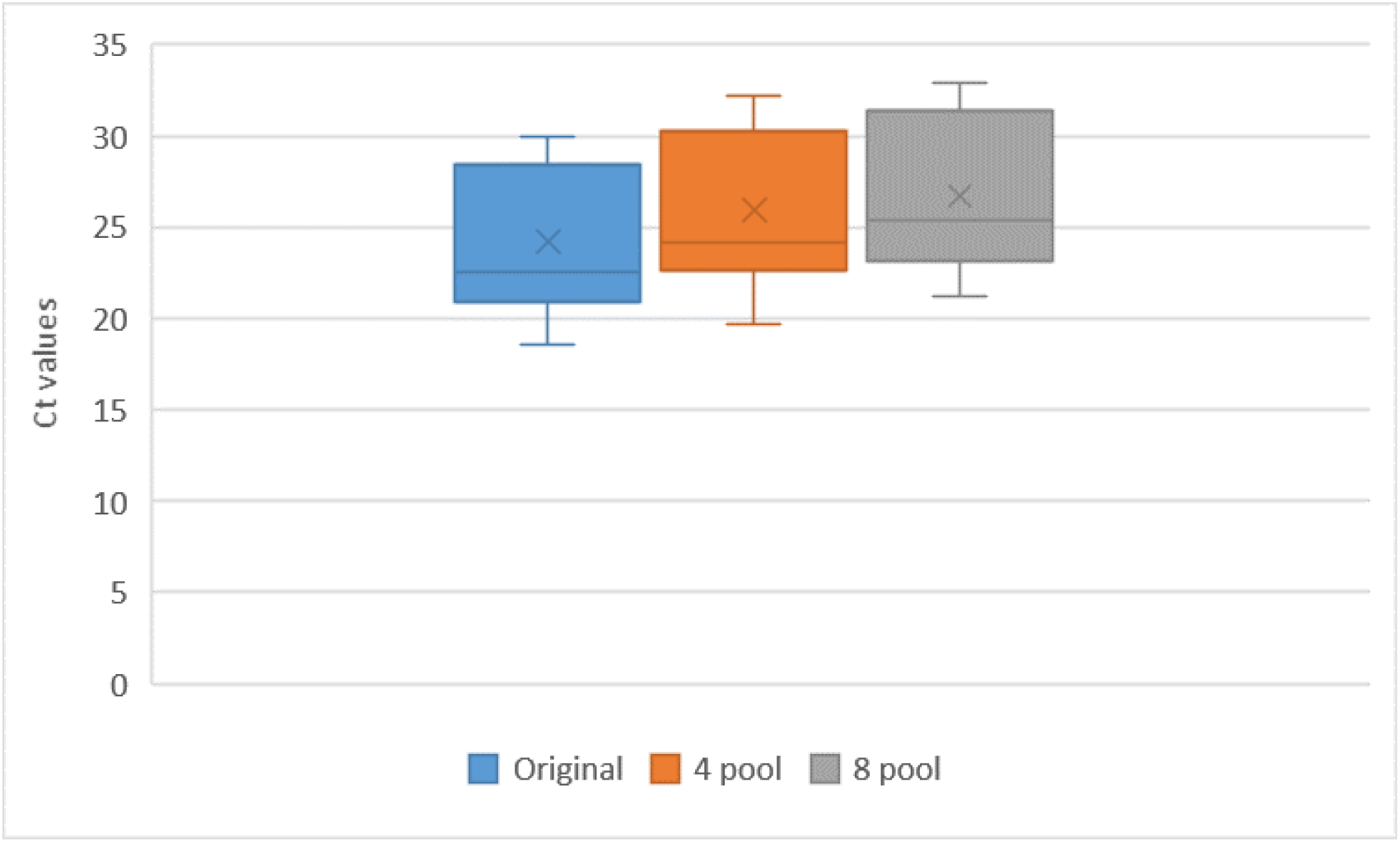
The Ct values of Medium viral load positive(MP) samples.

**Figure 4:**
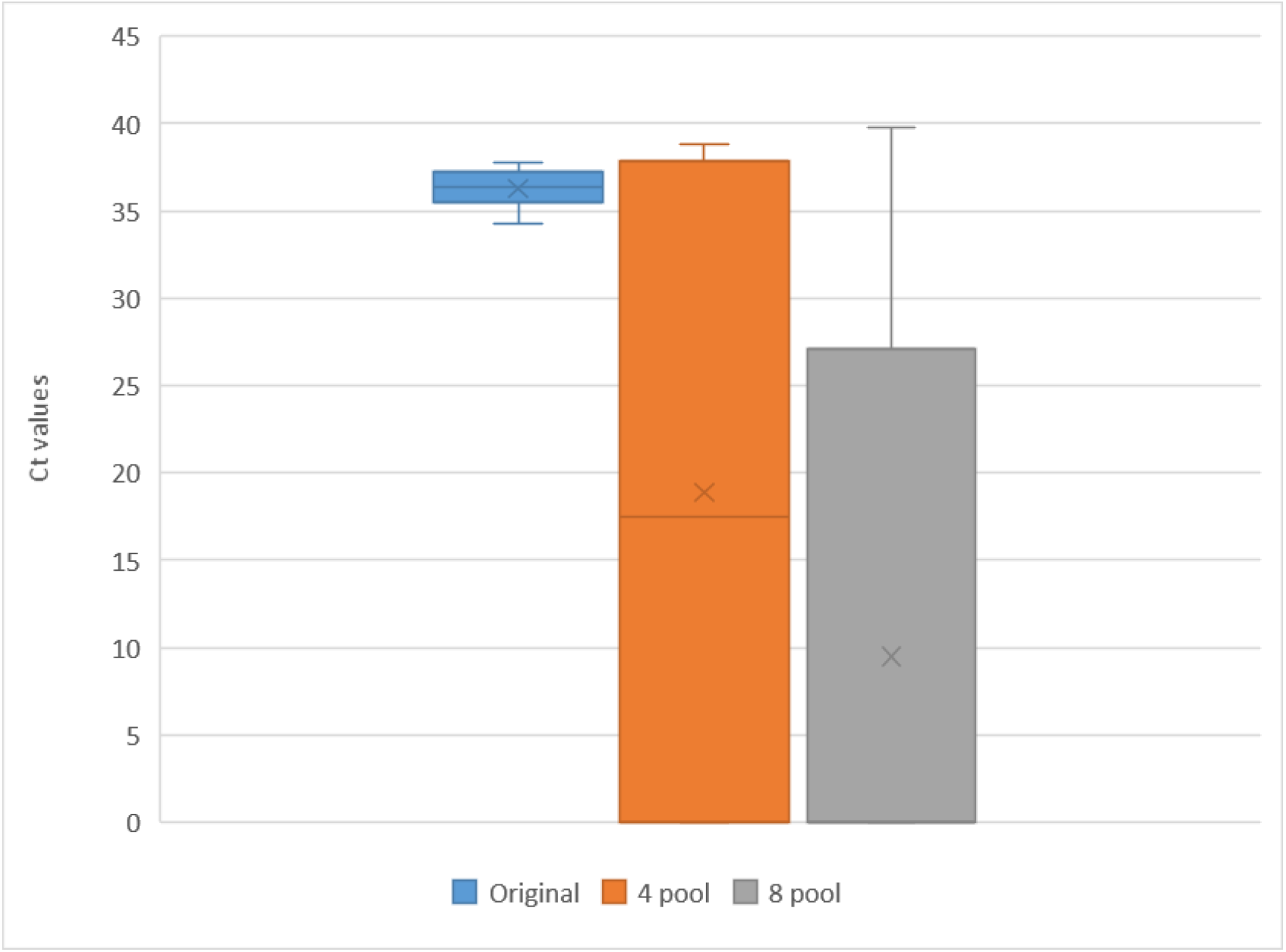
The Ct values of low viral load / weak positive (WP) samples.

Table 6, shows the discrepancies in results of the 40 positive samples, there were totally 15 positive samples that missed detection after the samples were pooled.

**Table 6:** Test result discrepancies in pool sample testing.

| Samples | Original | 4 pool testing | 8 pool testing |
| --- | --- | --- | --- |
| **High viral load positive samples (HP)** |  |  |  |
| 1 | Positive | Positive | Positive |
| 2 | Positive | Positive | Positive |
| 3 | Positive | Positive | Positive |
| 4 | Positive | Positive | Positive |
| 5 | Positive | Positive | Positive |
| 6 | Positive | Positive | Positive |
| 7 | Positive | Positive | Positive |
| 8 | Positive | Positive | Positive |
| 9 | Positive | Positive | Positive |
| 10 | Positive | Positive | Positive |
| **Medium viral load positive sample (MP)** |  |  |  |
| 11 | Positive | Positive | Positive |
| 12 | Positive | Positive | Positive |
| 13 | Positive | Positive | Positive |
| 14 | Positive | Positive | Positive |
| 15 | Positive | Positive | Positive |
| 16 | Positive | Positive | Positive |
| 17 | Positive | Positive | Positive |
| 18 | Positive | Positive | Positive |
| 19 | Positive | Positive | Positive |
| 20 | Positive | Positive | Positive |
| **Low viral load// weak positive samples (WP)** |  |  |  |
| 21 | Positive | Negative | Negative |
| 22 | Positive | Positive | Positive |
| 23 | Positive | Positive | Positive |
| 24 | Positive | Positive | Positive |
| 25 | Positive | Negative | Negative |
| 26 | Positive | Negative | Negative |
| 27 | Positive | Negative | Negative |
| 28 | Positive | Positive | Negative |
| 29 | Positive | Negative | Negative |
| 30 | Positive | Positive | Negative |
| 31 | Positive | Negative | Negative |
| 32 | Positive | Negative | Negative |
| 33 | Positive | Positive | Positive |
| 34 | Positive | Negative | Negative |
| 35 | Positive | Positive | Positive |
| 36 | Positive | Negative | Negative |
| 37 | Positive | Positive | Negative |
| 38 | Positive | Positive | Negative |
| 39 | Positive | Positive | Negative |
| 40 | Positive | Negative | Negative |

The % CV (Coefficient of variance) ranged from 0 – 4% in intra and inter assay of 4 pool samples of high and medium viral load positive samples, however inter assay of 8 pool sample and weak positive samples of 4 pool and 8 pool both inter and intra assay were very high (Table 7 and Table 8)

**Table 7:**
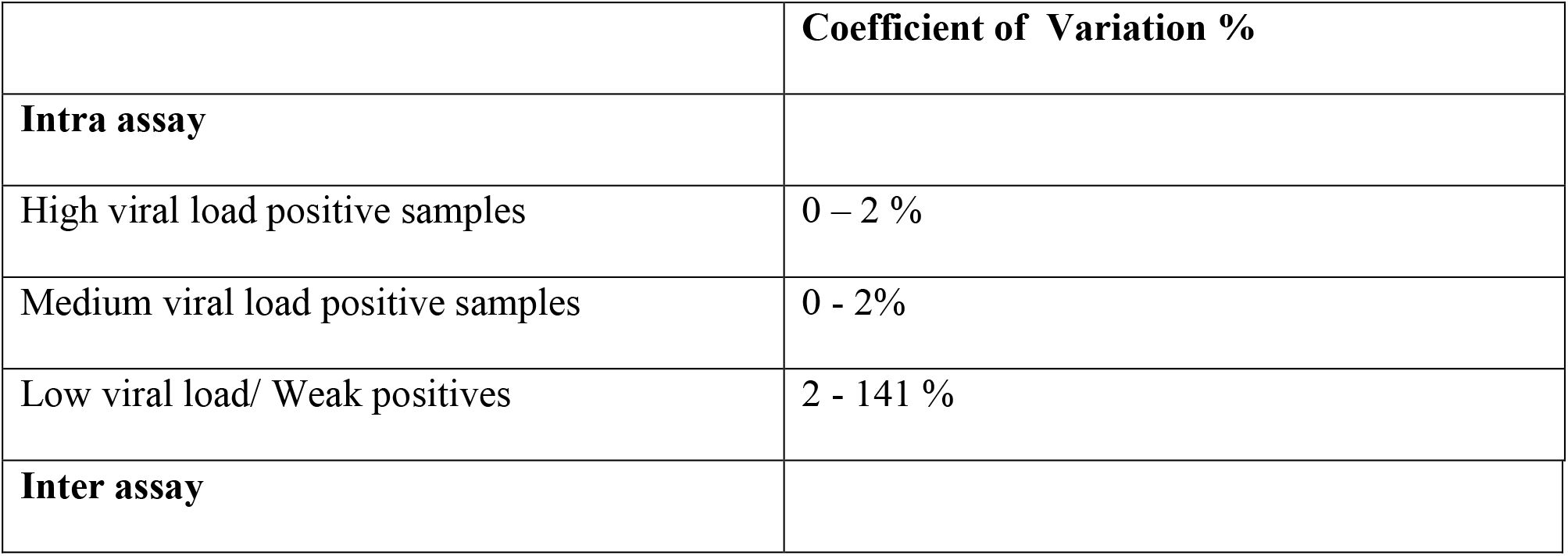
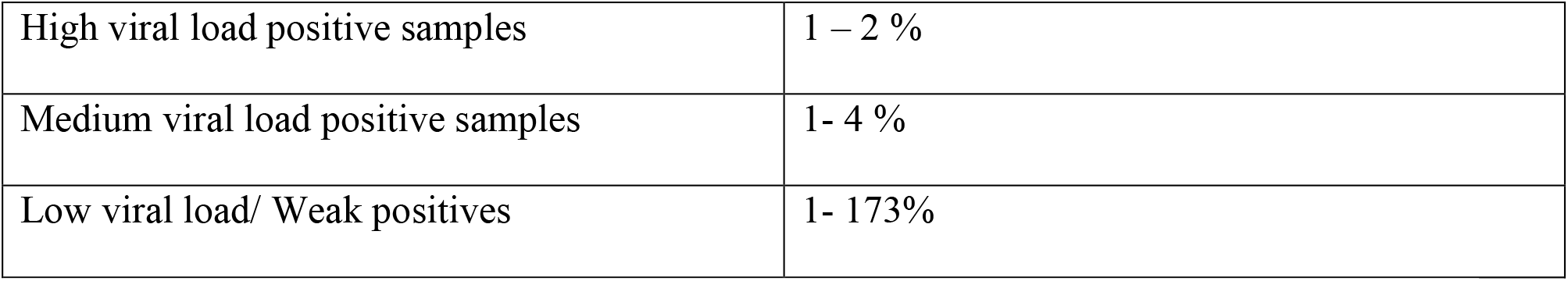
Intra and inter assay of 4 pooled sampling.

|  | Coefficient of Variation % |
| --- | --- |
| **Intra assay** |  |
| High viral load positive samples | 0 – 2 % |
| Medium viral load positive samples | 0 - 2% |
| Low viral load/ Weak positives | 2 - 141 % |
| **Inter assay** |  |
| High viral load positive samples | 1 – 2 % |
| Medium viral load positive samples | 1- 4 % |
| Low viral load/ Weak positives | 1- 173% |

**Table 8:**
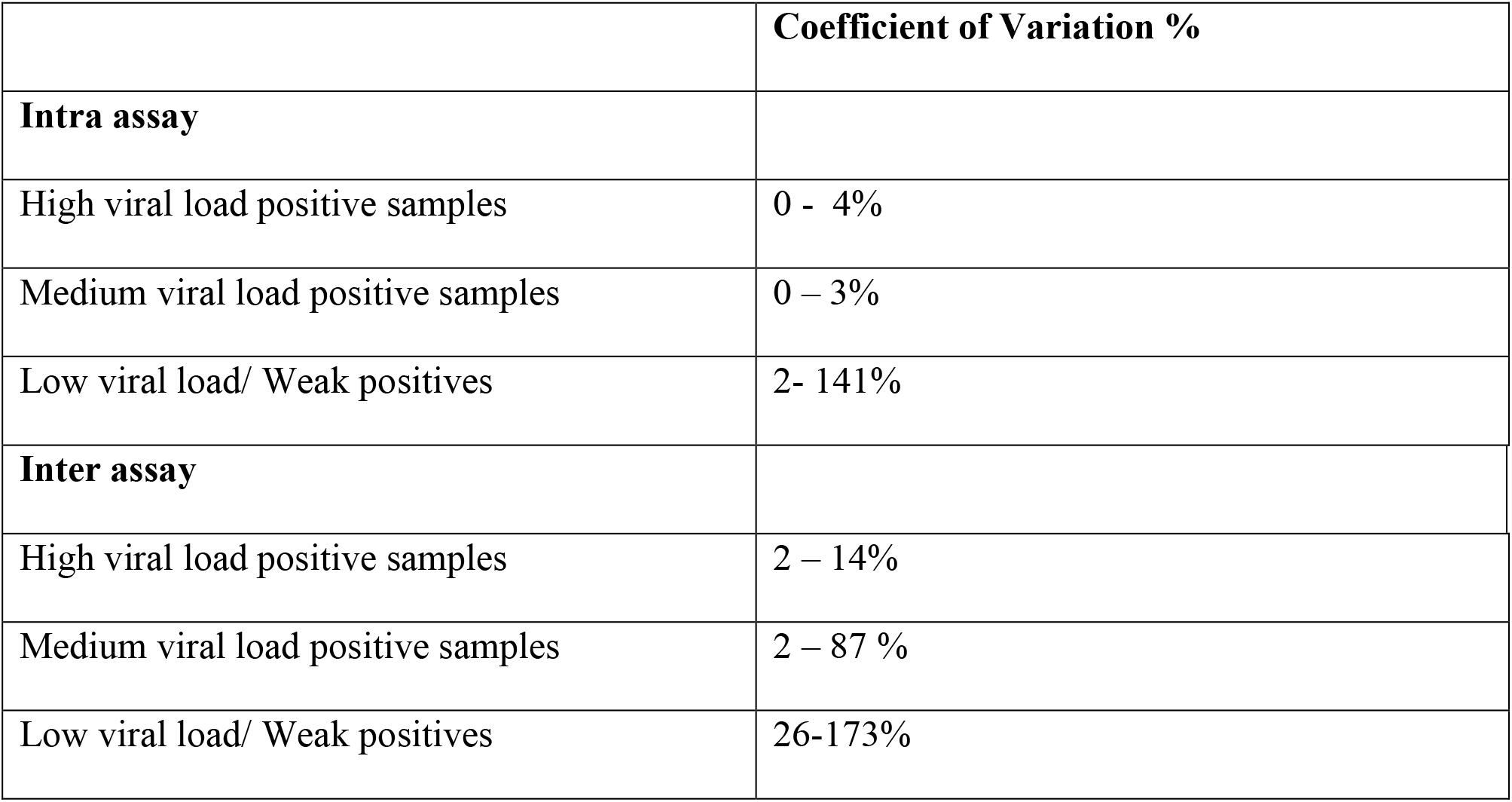
Intra and inter assay of 8 pooled sampling.

## Discussion

Our study showed that the strategy of pooling of samples for COVID-19 RT-PCR, has lower sensitivity than the standard individual RT-PCR and the sensitivity decreases with the increase in the pool size. Further when weak viral load positive samples were pooled the sensitivity became as low as 25%, which leads to increased false negatives. Similar studies have reported varied results, some studies have claimed pooling to be an effective strategy, a study done in Malaysia showed that pools of 10 samples were similar in sensitivity compared to individual testing. ^14^ A study in UK, has strongly supported pooled approach and reports a clinically insignificant sensitivity loss when samples are pooled and false positive rate up to 5.3%. ^15^ But another study done in Spain evaluating the sensitivity of pooled samples, testing various solutions and VTM showed that the sensitivity varied from 62.5 % to 81% and false negatives were as high as up to 40 %. ^16^

The study also showed that the pooling resulted in a median loss of 2.29 Ct for high viral load positive 4 -pool samples to median loss of – 36.01 Ct in 8 - pool low viral load positive samples, this is because the samples became negative with Ct value 0, after 8 sample pooling. This shows that in low viral load/ weak positive pooled samples the cycle threshold increased, or most samples went undetected. A study in Kenya also showed that the cycle threshold values were higher for samples which were pooled then tested individually. ^17^ The study in Spain also showed that sample dilution in pooling strategy resulted in a median loss of 2.8 to 3.3 Ct and thereby drop in sensitivity. ^18^

The coefficient of variation was high among the low viral load positive samples both in the inter and intra assay, this shows the high variability in the results when the samples are pooled. Higher Ct values in sample pooling method might be due to the inadequacy of samples when they are pooled and lower Ct values in pool testing might be due to the carrier effect of the higher RNA content in the pool. There is not much studies that have commented on %CV variability pertaining to COVID-19 testing. ^19^

When a person shows a weak positive result, that means that the viral load is low, so we need to amplify it several more times to detect the virus, hence the high Ct value. This kind of weak positives may be a of result of sample inadequacy or the person is in early/ late stage of infection where the viral load is low. These samples are missed detection when they are pooled. Such weak positive samples may pose a significant threat of spreading the infection to others. Studies on the clinical significance of low viral load positive cases are needed, and it remains a question of whether it is safe to miss detection of such cases?

Pooling samples can be very challenging logistically, usually in pool testing if a pooled sample tests negative, then all samples within that pool are deemed negative and if a pool tests positive, then individual samples making up that pool are tested again as individual samples to identify the positives. Hence the larger the pool the more challenging is the deconvolution. ^20^ Further there is additional time required in deconvoluting larger pools, which leads to time delay in reporting and this becomes crucial in severely ill patients and the high-risk contacts where early reporting is requisite. ^21^ Not to neglect the anxiety caused by isolation in patients included in the positive sample pools, until retesting of the individual samples is completed.

A lot of factors affect the sensitivity of RT-PCR, like the sensitivity of the kit, dilution used, the techniques of sample collection, type of samples (NPS vs oropharyngeal vs nasal etc), sample transport temperature, viral load in the sample which varies according to the stage of infection. ^22,23^ Therefore RT-PCR of sample pooling strategy has its own limitations, it cannot ensure the diagnostic integrity of the individual sample, ^24^ it can mask the technical errors like insufficient sampling, ^19,25^ it has higher percentage of false negatives, ^10^ reduces sensitivity ^26^ and sample dilution has led to the risk of missing weak viral load or borderline positive samples ^17,21^ Although pooling of samples facilitates rational use of resources, it might miss individuals who might be positive for COVID-19. ^27,28^

This information becomes crucial for deciding the pooled strategy testing for disease like COVID −19. In the current pandemic, we simply can’t afford to miss any positive cases. With the high Ro value around 3 for SARS-COV-2, ^29^ every missed positive case can lead to rapid spread of infection to others.

## Limitations

Our study was conducted in a single laboratory in a defined geographical area, more studies and data are needed to validate sample pooling strategies and the clinical significance and threat of communicability of weak positive samples.

## Conclusion

Based on the study results, we conclude that pooled testing strategy misses low viral load/ weak positive COVID – 19 cases and there is high variability in results when the samples are pooled. While in a low resource setting, where pooled sample testing is better than not having any kind of testing, sample pooling might be an effective way for mass screening and the increase in percentage of false negative tests may be dismissed, but caution is advised and it needs careful scrutiny. In the current scenario of the pandemic, validation studies on the cost-effective analysis of pooling samples should be done, considering the cost and risk of missing even a single positive person. As COVID - 19 numbers are still high and testing capacity needs to be high, we have to meticulously evaluate the testing strategy of each country depending on its testing capacity, infrastructure, economic strength, and need to make a serious call on cost effective strategy of resource saving and cost of missing positive patients.

## Data Availability

The data is available with the corresponding author, Dr. Sally, Director of Biogenix G42 lab and will be produced on request

## Declarations

### Ethics approval and consent to participate

The Ethics approval was obtained from Department of Health (DOH) Institutional review board (IRB), Abu Dhabi

All methods were carried out in accordance with relevant guidelines and regulations. The Department Of Health (DOH) Institutional review board (IRB), Abu Dhabi has waived off the consent as it does not use any participant data/ information, the lab data on the viral samples were only used for this study

Consent for publication – Not applicable

### Competing interests

The authors declare that they have no competing interests regarding the publication of this paper

### Funding statement

The study was not funded by any funding body, it was done in Biogenix lab as a part of research

### Authors’ contribution

SM – conception, design of work, acquisition, analysis, interpretation of data, drafted and substantively revised the manuscript

EI – conception, design of work, acquisition, analysis, interpretation of data, drafted and substantively revised the manuscript

BT- conception, design of work, interpretation of data, drafted and substantively revised the manuscript

JT- conception, design of work, interpretation of data, drafted and substantively revised the manuscript

PR- conception, design of work, drafted and substantively revised the manuscript

SG- Acquisition, analysis, interpretation of data, drafted and substantively revised the manuscript

WA- conception, drafted and substantively revised the manuscript

## Acknowledgments

We acknowledge all lab personnel, skilled technicians who provided help during the research and preparation of the manuscript

## Notes

### Competing Interest Statement

The authors have declared no competing interest.

### Author Declarations

The Ethics approval was obtained from Department of Health (DOH) Institutional review board (IRB), Abu Dhabi All methods were carried out in accordance with relevant guidelines and regulations. The Department Of Health (DOH) Institutional review board (IRB), Abu Dhabi has waived off the consent as it does not use any participant data/ information, the lab data on the viral samples were only used for this study

